# An Implementation Science Framework to Understand Low Coverage in Mass Dog Rabies Vaccination

**DOI:** 10.1101/2025.02.05.25321168

**Authors:** Ricardo Castillo-Neyra, Lizzie Ortiz-Cam, Jorge L. Cañari-Casaño, Elvis W. Diaz, Laura D. Tamayo, Guillermo Porras, Sergio E. Recuenco, Valerie A. Paz-Soldan

## Abstract

**Background:** Dog-mediated human rabies has been greatly reduced in the Americas and eliminated from most high-income countries. However, many countries in Africa, Asia, and parts of Latin America are still struggling with this gruesome disease. Mass dog vaccination, a One Health strategy, is the primary approach for elimination. However, achieving and sustaining appropriate vaccination coverage in endemic areas remains a challenge. Our objective was to apply the Consolidated Framework for Implementation Research (CFIR) in Arequipa, Peru as a guiding tool to understand the barriers faced by different stakeholders.

**Methods:** Seven focus groups with 56 participants were conducted to capture community perspectives on rabies and vaccination. A workshop was conducted with two groups of public health personnel (n= 69): mass dog vaccination campaign (MDVC) implementers and authorities, in charge of dog rabies control. With these stakeholders we explored factors contributing to the decrease in MDVC post COVID-19. We used the CFIR approach to understand barriers within five different domains: innovation, outer setting, inner setting, individuals, and implementation.

**Findings:** Barriers within the community included insufficient communication, a short vaccination schedule, and fragmented collaboration between system coordinators. At the individual level, a decreased perception of rabies risk occurred as both people and their dogs spent more time indoors due to the COVID-19 pandemic (in urban areas). Dog vaccination was deprioritized compared to COVID protection, with individuals focusing on their own vaccinations and avoiding crowded spaces. In peri-urban areas, longer work hours due to the pandemic’s financial impact left less time for dog vaccinations on weekends. Participants reported confusion caused by private veterinarians, who claimed that government-subsidized vaccines were of poor quality. Among implementers and authorities, the main barriers included insufficient MDVC materials and equipment, unclear responsibilities, and a lack of time to evaluate the campaign after activities. Importantly, financial constraints and fragmented commitment from higher-level institutions posed challenges for proper planning and implementation.

**Conclusions:** We identified barriers and co-designed strategies to improve MDVC participation including strengthening municipal alliances, virtual and physical publicity for events within districts, adequate training for vaccinators, reinforcing vaccinators to remain in fixed spots, and expanding vaccination campaign hours.

**Author summary:** Dog-mediated human rabies, a fatal and preventable disease, remains a significant public health challenge in some parts of Latin America, including Arequipa, Peru. Despite efforts to control the disease through mass dog vaccination campaigns (MDVCs), achieving and maintaining adequate vaccination coverage has proven difficult, especially in the aftermath of the COVID-19 pandemic.

To address this issue, we applied the Consolidated Framework for Implementation Research (CFIR), a widely used tool in implementation science, to identify barriers to MDVC implementation and propose actionable solutions. Through focus groups with community members and a workshop with public health personnel, we identified key challenges, including fragmented communication, decreased public perception of rabies risk, financial and logistical constraints, and misinformation about vaccine quality. Our findings reveal that the pandemic not only disrupted vaccination campaigns but also shifted public and institutional priorities away from rabies prevention. By engaging with stakeholders, we co-designed strategies to overcome these barriers, such as improving interinstitutional collaboration, enhancing public awareness, and extending campaign hours to better accommodate the community’s needs. This study underscores the importance of a One Health approach to understand the complexities of controlling a canine rabies outbreak, including integrating perspectives from diverse stakeholders to develop sustainable solutions for rabies control. Our recommendations aim to strengthen rabies elimination efforts in Arequipa and can inform similar strategies in other endemic regions.

## Introduction

Dog-mediated human rabies remains a public health problem in some Latin American countries^1^. Mass dog vaccination is an evidence-based and cost-effective intervention to control and eliminate dog and dog-mediated human rabies^2–4^. Achieving herd immunity requires vaccinating at least 70% of the dog population at risk^2^, with the Pan American Health Organization recommending a minimum of 80%^4^ coverage for the Americas. The city of Arequipa, Peru is in the middle of a dog rabies epidemic that was first detected in March 2015^5^. Alarmingly, despite the proven effectiveness of mass dog vaccination campaigns (MDVCs) in other Latin American countries, the number of rabid dogs in Arequipa continues to increase, and vaccination coverage has not only remained below target but has also declined in recent years^6–8^.

MDVCs are true One Health interventions, focusing exclusively on vaccinating domestic dogs—the animal reservoir—to protect human health. MDVCs typically operate on fixed vaccination points in central locations for a single or few days^9^. Some studies suggest using data-driven algorithms to optimize resource allocation and enhance vaccination participation and equity^10,11^. In Peru, MDVCs employ three strategies based on the setting: fixed-point vaccination, door-to-door vaccination, and mobile teams rotating through various locations^12^. Fixed vaccination sites are usually chosen for their convenience and prominence, such as well-known parks^12^. Despite these efforts, vaccination coverage in Arequipa has been declining. The MDVC was carefully planned with local authorities and implementers to ensure a clear and cohesive plan, but field monitoring revealed shortcomings, particularly a lack of adherence to established protocols and plans. Other factors, such as the COVID-19 pandemic further exacerbated the situation, diverting health authorities’ focus and resources, leading to reduced or canceled MDVCs^13^, which in turn contributed to an increase in cases and a geographic expansion of the outbreak to new areas^14^. Additionally, community fears of contracting COVID-19, a lack of timely and accurate information about MDVC logistics, and insufficient planning and resources have hindered these efforts^8^.

Implementation science is crucial for identifying and addressing barriers that hinder the adoption of evidence-based interventions in public health programs. This discipline has been instrumental in refining and adapting strategies for specific contexts – such as that for measles vaccination^15^, and HIV and tuberculosis control^16,17^ – as well as enhancing community acceptance and adherence to treatment through educational campaigns, social support, and technological innovations. The Consolidated Framework for Implementation Research (CFIR) is highly regarded as an optimal framework for guiding the implementation of practices and interventions, due to its comprehensive and structured approach (covering numerous domains), ability to consider various contextual factors, and flexibility for both prospective and retrospective use^14^.

The main objective of this study was to examine the multilevel factors contributing to the low levels in MDVC coverage during a dog rabies epidemic, both 1) generally (i.e., pre-pandemic), and 2) post COVID-19. We used the CFIR to organize the range of responses received from the different stakeholders using specific domains to address intricate, interrelated, and multilevel ideas^18^ to pinpoint barriers that impede adequate vaccination coverage.

## Methods

### Study Setting

Arequipa City has 1,031,727 inhabitants^19^ and is located at 2,300 meters above sea level. The city has 14 urban and periurban districts^20^ with diverse socioeconomic status (SES). Emerging periurban neighborhoods around the city are inhabited mostly by migrants with lower SES than member of well-established urban communities. When these periurban neighborhoods urbanize they witness an improvement in connectivity with the broader city, marked by the development of improved sidewalks, road infrastructure, and enhanced transportation access^21,22^. In this city, the rabies virus has been circulating in the dog population at least since March 2015, when the first detected rabid dog was reported^5,23^. Since then, additional rabid dogs have been identified almost weekly in previous years. In 2024, detection of cases dropped, but also the submission of samples. By September 2024, there had been 392 confirmed cases of rabid dogs and one human case^24–26^.

### Study Design

Our research team had been conducting Chagas research in Arequipa for over a decade when the rabies outbreak started, and quickly shifted and obtained funding to study this new outbreak, led by a male veterinarian and PhD in epidemiology (RCN), a female social scientist with ample experience in qualitative research with a PhD in public health (VPS), and a dedicated male veterinarian to manage the study (EWD). Once we identified a decrease in MDVC participation, we triangulated methods and data sources to obtain a range of perspectives about the potential factors contributing to this decrease. We conducted seven focus groups to obtain community perspectives on barriers to their MDVC participation, during and after the COVID-19 peak. We also facilitated fifteen small focus groups with MDVC implementers and authorities to obtain their perspectives on barriers and facilitators for mass dog vaccination, conducted during a day-long group model building^27,28^ workshop, aimed at planning for an improved MDVC. Finally, we examined our research team’s field notes of observations during MDVCs.

### Study Population

We had three main study populations.

- Community members: Adult women from urban and periurban areas within Arequipa city. Based on surveys and previous focus groups, women are the ones who mostly decide on the vaccination of dogs and have more knowledge about the home’s dog. All participants had to be dog owners and had no prior relationship with the research team.
- MDVC implementers: Health personnel from *microreds (micro-health networks)*, which are the health system operational units that are about the size of a district (~10,000 households). These implementers are health inspectors, public health veterinarians, cold chain managers, health promotion managers, and communication managers. Our research team has been collaborating with MDVC implementers in this region for about two decades – first, for studies related to Chagas disease, and since 2015, when rabies outbreak started. Our study team did not have a prior relationship with the vaccinators who are volunteers.
- MDVC authorities: Key personnel from the two public entities overseeing the healthcare system: the “Gerencia Regional de Salud of Arequipa”, which serves as the regulatory body and equivalent of a regional Ministry of Health, and the “Red de Salud Arequipa-Caylloma”, the operational entity responsible for managing the *microreds*. These authorities dictate the local guidelines, request budgets, approve *microred’s* plans, provide technical support for the activities, plan the city-wide health programs, manage and submit data to the national level, and monitor the program’s activities. These authorities include regional coordinators of the various strategies: zoonotic diseases, immunization (including cold chain), health promotion, and communication. The authorities also included regional laboratory coordinators for rabies diagnosis, and representatives from both the epidemiology regional office, and the human health division. Our research team has been collaborating with the MDVC authorities for several years.

Additionally, our research field team members (EWD, GP, and/or JC) visited *microreds* before and during the MDVCs and provided detailed field notes on the planning and implementation processes of MDVCs conducted in 2022.

### Sample and Recruitment

Our research team used purposive sampling to recruit 56 community members for focus groups through a two-phase process, selecting women who owned dogs from four urban and three periurban areas from a randomly selected area of about nine neighborhood blocks that were at least 5 blocks away from a health facility to minimize potential bias from living near a health facility and being more exposed to health-related information. We conducted door-to-door (face-to-face) recruitment two hours before the event, enlisting 25 participants from periurban and 31 from urban areas. We exclusively recruited women who owned dogs but had not participated in the last MDVC. These women ranged in age from 18 to 76, with a median age of 40. After achieving theme saturation in five focus groups, we conducted two additional groups to confirm no new topics emerged.

We organized an in-person daylong workshop in Arequipa, inviting 80 MDVC implementers and authorities. During this workshop, attended by 69 participants, we conducted 14 focus groups with implementers: 13 with four implementers each and one with six implementers. These participants represented 20 different *microreds*. We randomized implementers into each group to ensure diversity and enhance discussion. Additionally, we held a focus group with the 11 authorities attending the workshop. This workshop aimed to present monitoring data, reflect on findings, and discuss improvements for future MDVC planning processes.

Finally, during each 2022 MDVC in Arequipa, at least one member of our research team actively observed the vaccination activities and took field notes on implementation processes and fidelity to plans established *a priori*. Most of the time, they also attended the *microred* offices during the MDVC planning sessions.

### Data Collection Tools and Processes

The research team developed a focus group guide to ensure similar questions were asked of each focus group to explore the main topic of interest: the factors at individual, community, and systems levels that contributed to the decrease in MDVC coverage post COVID-19. We also explored related topics, including knowledge about rabies, rabies vaccination campaigns, and changes in the free-roaming dog population and dog tenure during and after the peak of the COVID-19 pandemic. The team included researchers with extensive experience in qualitative methods and public health, specifically in the context of zoonotic diseases: a social scientist as facilitator (VPS), an epidemiologist (RCN), and a veterinarian (EWD), and a research associate (Peruvian epidemiology doctoral student at the time) taking notes (JC).

Before each focus group, the facilitator made aware of the study’s objectives, including understanding the barriers to mass dog rabies vaccination (MDVCs) and reviewed the consent form with participants and obtained written consent regarding voluntary participation and agreeing to audio recording. We also emphasized at the start of each focus group the importance of obtaining different points of view and that there were no correct or incorrect answers to ensure that participants felt free to share their views without others’ influence. Focus groups were conducted in schools or courtyards, without the presence of onlookers or non-participants. The focus groups lasted around 60-90 minutes.

The workshop with MDVC implementers and authorities was also led by an experienced social scientist as facilitator (VPS) and rabies content expert (RCN), with fourteen co-facilitators from various fields (veterinarians, psychologists, biologists, political scientists, and administrators). Here, the facilitators informed the participants about the study’s objectives and obtained written consent for participation and audio recording. The workshop began with a short sharing of preliminary findings of MDVC coverage in the different regions (anonymized to prevent judgement) and then proceeded to break out focus groups where data was collected. No repeat focus groups were conducted. Only invitees were present.

For the MDVC observations, our rabies research manager (EWD) observed 41 MDVCs in action, from June to October 2022. He recorded notes and observations on a google document that was shared between him and the project PI (RCN) and discussed with the team at weekly meetings.

### Data Processing and Analysis

After each community-based focus group, the research team discussed and summarized the themes that emerged. Once they conducted the last focus group, the team met to discuss the overall themes that had emerged inductively in preparation for the development of the codebook. After finalizing the workshop with MDVC implementers and authorities, the research team discussed new themes to add to the same codebook. The research team used a mix of inductive and deductive codes: CFIR domains and subdomains were used to organize data for analysis (deductive), using themes that emerged during data collection (inductive). All recordings were transcribed and checked for quality.

Between March and May 2023, using the codebook, the research team (LOC, JC) analyzed the double-transcripts using Dedoose version 9.0.90 for all focus group, workshop, and field note data (which was already in a google document as noted by the observer (EWD)). They resolved discrepancies through discussions and then proceeded to code additional transcripts. During the analysis, findings from community focus groups were stratified by setting type: urban versus periurban. All data was analyzed without returning transcripts to participants (whether community or MDVC implementers).

## Results

We mapped results from focus groups with community members, MDVC implementers, authorities, and observation field notes (Research Field Team) using the updated CFIR^18^. We organized the data within the five CFIR domains (Table 1) according to the number of barriers identified in each domain. Most barriers appeared in the Implementation Process domain, followed by the Inner, Individuals, and Outer Setting domains. The Innovation domain contained the fewest barriers.

**Table 1.**
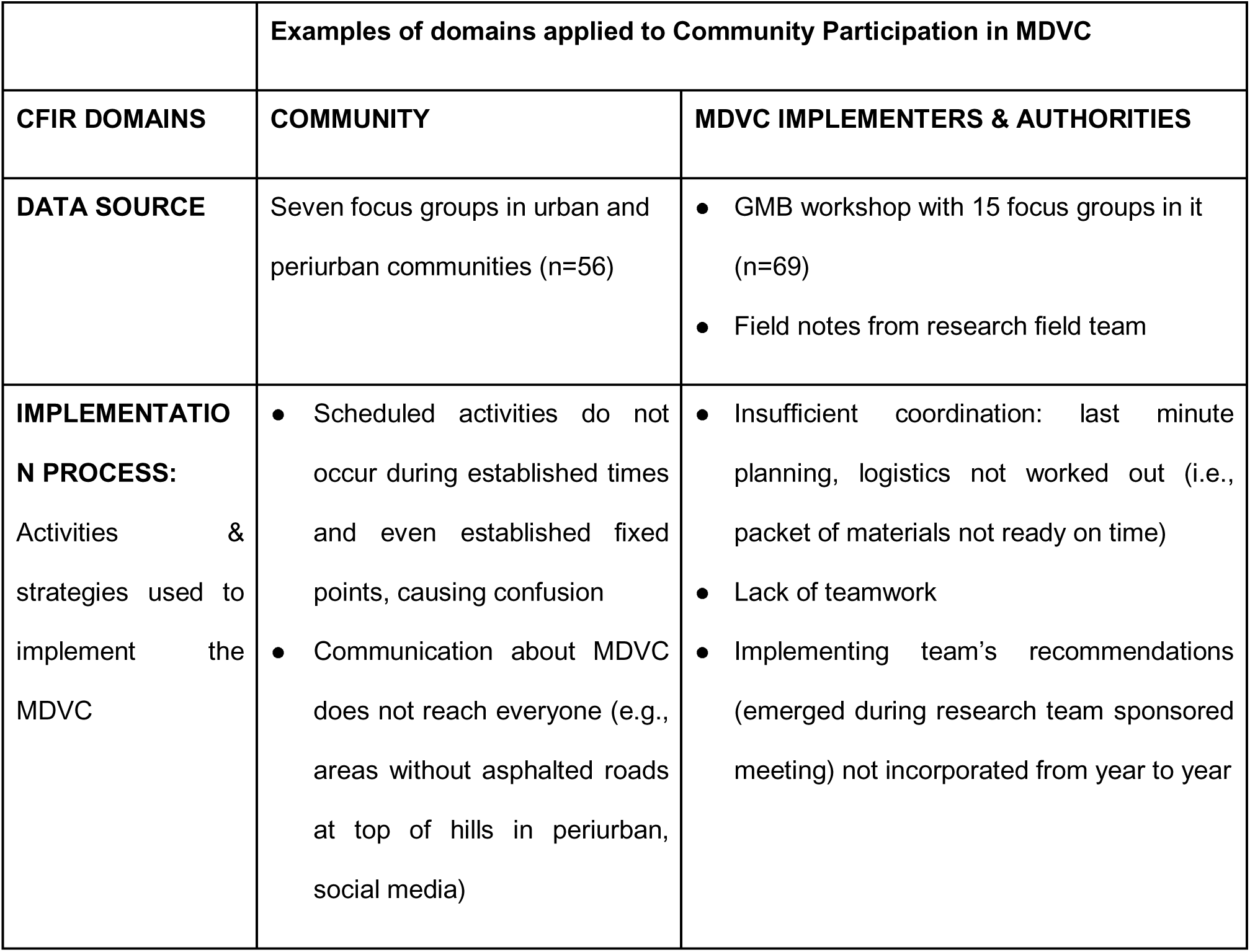
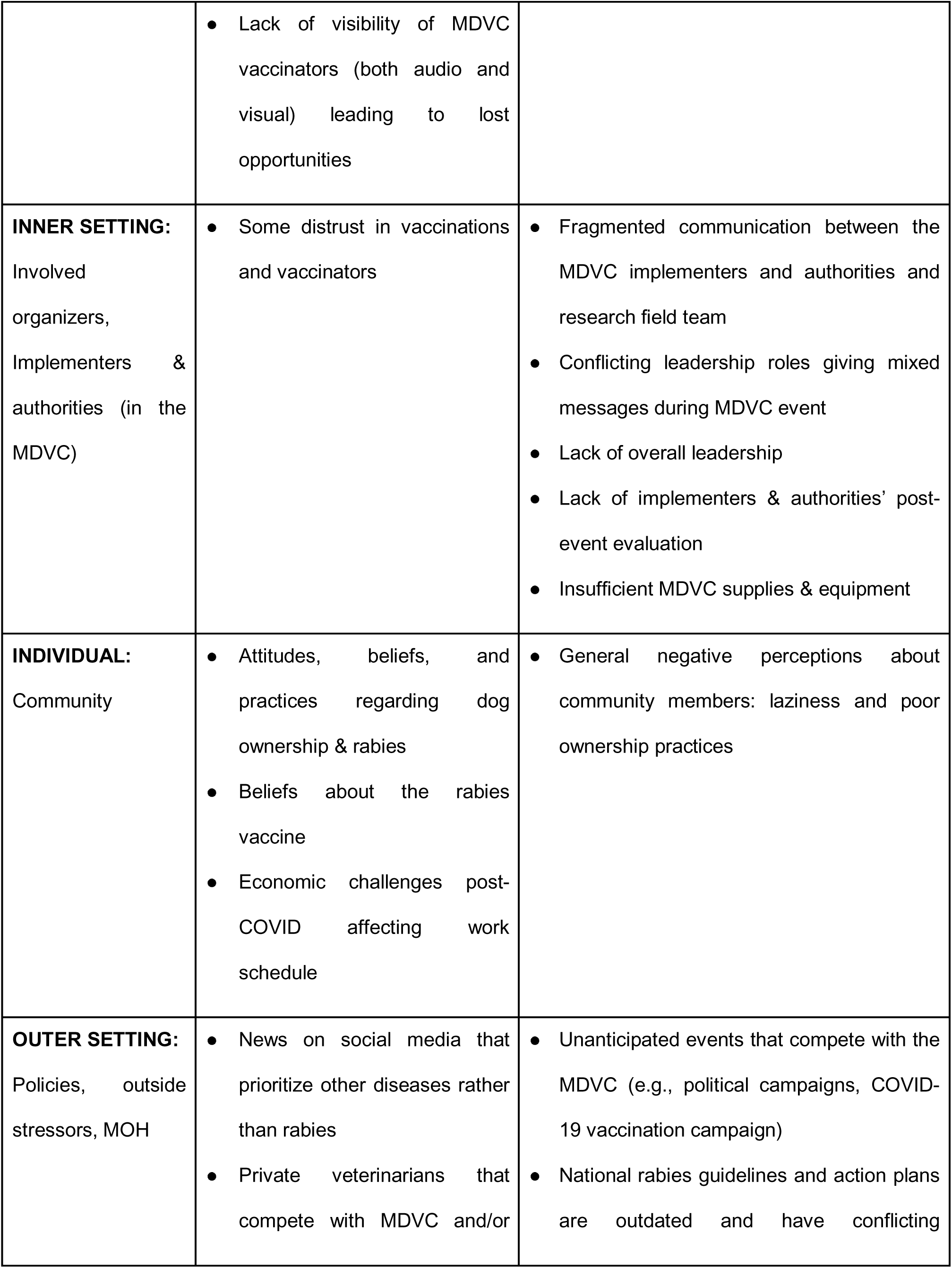
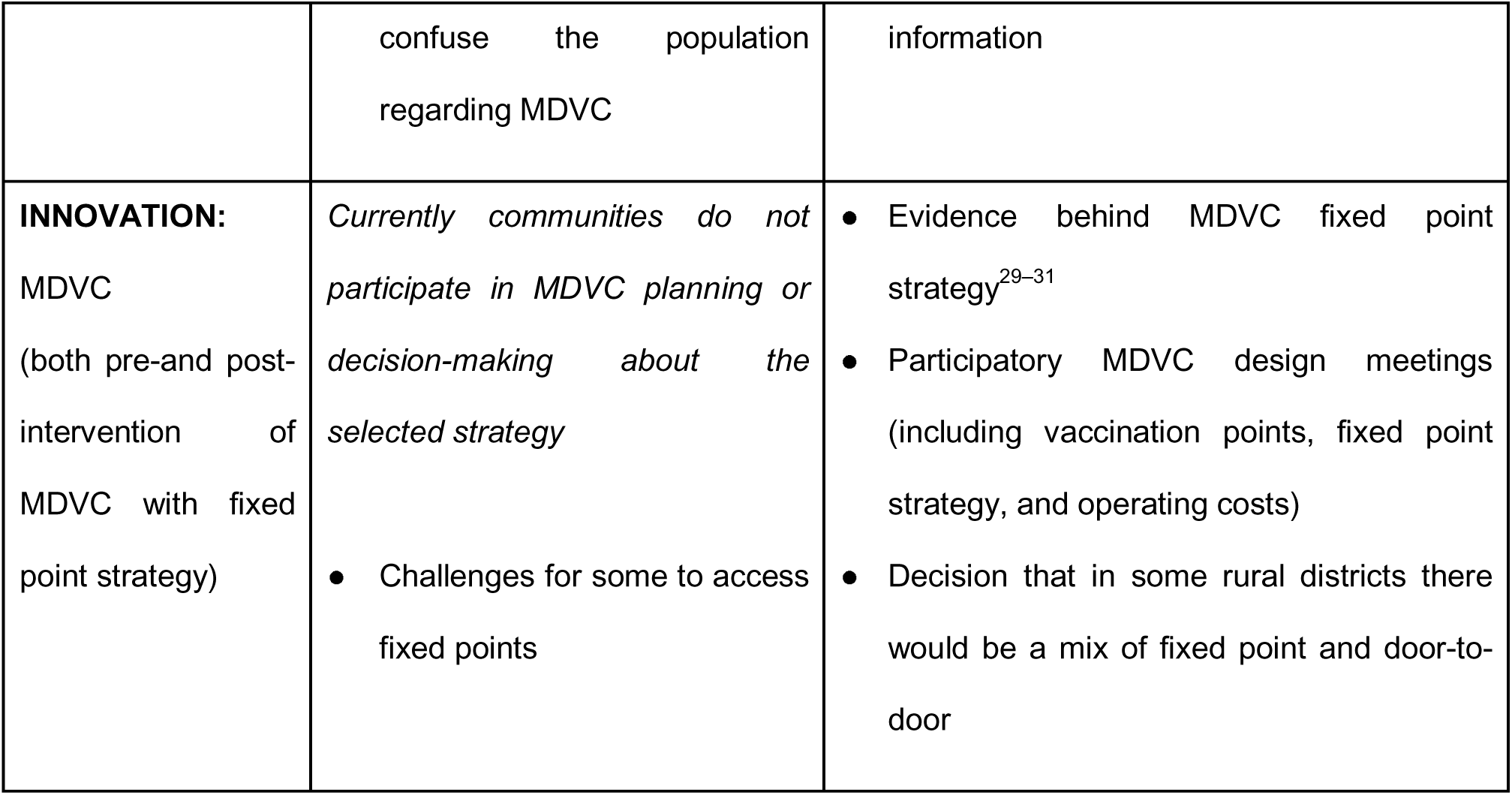
Examples of CFIR domains adapted to our research.

|  | **Examples of domains applied to Community Participation in MDVC** |  |
| --- | --- | --- |
| **CFIR DOMAINS** | **COMMUNITY** | **MDVC IMPLEMENTERS & AUTHORITIES** |
| **DATA SOURCE** | Seven focus groups in urban and periurban communities (n=56) | • GMB workshop with 15 focus groups in it (n=69)<br>• Field notes from research field team |
| **IMPLEMENTATION PROCESS:**<br>Activities & strategies used to implement the MDVC | • Scheduled activities do not occur during established times and even established fixed points, causing confusion<br>• Communication about MDVC does not reach everyone (e.g., areas without asphalted roads at top of hills in periurban, social media) | • Insufficient coordination: last minute planning, logistics not worked out (i.e., packet of materials not ready on time)<br>• Lack of teamwork<br>• Implementing team's recommendations (emerged during research team sponsored meeting) not incorporated from year to year |
|  | • Lack of visibility of MDVC vaccinators (both audio and visual) leading to lost opportunities |  |
| **INNER SETTING:**<br>Involved organizers, Implementers & authorities (in the MDVC) | • Some distrust in vaccinations and vaccinators | • Fragmented communication between the MDVC implementers and authorities and research field team<br>• Conflicting leadership roles giving mixed messages during MDVC event<br>• Lack of overall leadership<br>• Lack of implementers & authorities' post-event evaluation<br>• Insufficient MDVC supplies & equipment |
| **INDIVIDUAL:**<br>Community | • Attitudes, beliefs, and practices regarding dog ownership & rabies<br>• Beliefs about the rabies vaccine<br>• Economic challenges post-COVID affecting work schedule | • General negative perceptions about community members: laziness and poor ownership practices |
| **OUTER SETTING:**<br>Policies, outside stressors, MOH | • News on social media that prioritize other diseases rather than rabies<br>• Private veterinarians that compete with MDVC and/or | • Unanticipated events that compete with the MDVC (e.g., political campaigns, COVID-19 vaccination campaign)<br>• National rabies guidelines and action plans are outdated and have conflicting |
|  | confuse the population<br>regarding MDVC | information |
| **INNOVATION:**<br>MDVC<br>(both pre-and post-intervention of MDVC with fixed point strategy) | *Currently communities do not participate in MDVC planning or decision-making about the selected strategy*<br>Challenges for some to access fixed points | Evidence behind MDVC fixed point strategy<sup>29–31</sup><br>Participatory MDVC design meetings (including vaccination points, fixed point strategy, and operating costs)<br>Decision that in some rural districts there would be a mix of fixed point and door-to-door |

### Implementation Processes

Among community members, the most discussed barrier was poor communication about the dates and locations of the MDVC. In contrast, implementers and authorities frequently mentioned the need to establish adequate policies and laws to improve collaboration with municipalities as a major barrier. Community participants also noted inadequate information about upcoming events, such as learning about the vaccination campaign only when it was already in their neighborhood without time to plan. Additionally, they mentioned the limited scheduled time at fixed vaccination points, often finding vaccinators packing up to leave just as they arrived with their dogs.

> “There is not enough publicity like before when they used to stop at every corner: ‘Don’t forget this Sunday!’ and the diffusion of the campaign was from Monday to Friday, so you already had the chance to have a plan: ‘Oh, this Sunday you have to take Rocky, right?’ (Urban Community Member)

> “They used to put up posters, then they went from street to street during the whole week announcing when the campaign was going to take place” (Urban Community Member)

The MDVC implementers and authorities also mentioned using posters/banners and health promotion activities to create awareness in different areas. Nevertheless, community members describe that there was a lack of visible banners or, simply, of visibility overall about the MDVC:

> “When I saw that they were vaccinating, it was just a little table with a cooler, 2 people, that’s it. There was not even a banner or something that said ‘We are from the Ministry of Health’. I had to go and ask: ‘’What do you do here, do you vaccinate?” (Urban Community Member)

Community members suggested using a specific song associated with previous MDVC events and placing megaphones on garbage trucks that reach the most remote areas of different communities. Participants also recommended increasing physical publicity within their districts (i.e., schools, markets, bus stops), as well as virtual formats (i.e., social networks, radio) to allow the youngest members of the house to be engaged in this activity, since many times they are the ones who spend the most time using social media, being with their dogs, and being most attentive to their dogs’ needs:

> “… they [my girls] tell me ‘Coco is missing its vaccine shot’” (Urban Community Member)

> “…Sometimes they [the children] find out about the campaign when they come home from school and say ‘they are vaccinating, I’m going to bring my dog’… ‘They are vaccinating, they are vaccinating’, they shout” (Periurban Community Member)

Participants suggested adding dog deworming to the MDVC to add community interest: *‘I am sure that the movement of people would be greater’ (Urban Community Member)*

Community members and the research field team expressed concerns about the accessibility and visibility of fixed vaccination points, noting that locations were often unclear, far from homes, or poorly communicated, hindering access. The team also emphasized the need for vaccinators to remain at their posts longer to increase awareness. Additionally, community members recommended scheduling vaccination campaigns in the afternoons to accommodate those with morning work commitments.

> “The lady who has come to vaccinate has gone to the 7A bus stop, behind a little tree. She was hiding there with her vaccines. Whenever they have come to vaccinate, they have always come here to the soccer field” (Periurban Community Member)

> “… said that the vaccinator team is not walking, they are in places with poor visibility and that they do not turn on the megaphone in some cases” (Research field team)

> “The vaccine may be free, but it is so far that they [my neighbors] say… ‘I’d better pay my five soles here nearby and I don’t have to spend money on a cab’” (Urban Community Member)

> “I have seen the pasted ads on the buses, but it [the ad] says ‘Villa El Mirador’ and this is not Villa El Mirador, it is the ‘Mirador of Arequipa Independencia’, isn’t it?” (Periurban Community Member)

Likewise, the community and the research field team perceived that the vaccinators were not responsible nor committed to the purpose of the campaign:

> “… flyers were handed out and everything, but they didn’t come. They came at about 12 o’clock…” (Periurban Community Member)

> “Yes, they came here for a little while, half an hour, yes, half an hour exaggerating, half an hour, and from there I returned with my dogs, ‘no, we are leaving!’ They told me and left” (Periurban Community Member)

> “We went and […] they were already putting away their banners and I told them I was coming for the vaccination campaign, ‘no it’s over!’” (Urban Community Member)

> “The vaccination started at 3:20 pm and the delay of the registrar…. was a problem as I had to take her place to move forward with the vaccination since she arrived at 4:15 pm” (Research field team)

Some community members recommended using megaphones and social media platforms like Facebook to announce MDVC dates and locations. However, they also highlighted communication challenges with these methods. Megaphone announcements were often inaudible and limited to main roads, causing residents in peripheral areas to miss them. Additionally, one participant questioned the effectiveness of social media communication.

> “And the information is now all through Facebook, all virtual. So it no longer reaches the ears of the mothers who are at home and the children do not inform either” (Urban Community Member)

Additionally, field notes from the research team highlighted the need for coordination between MDVC implementers and authorities to ensure adequate logistical support and improve the efficiency and effectiveness of the vaccination campaign.

> “Then we had to wait for the mobility since [the municipality team] arrived late, so we started the vaccination late” (Research field Team)

> “The MDVC transport and megaphone vehicle [are] deficient” (Research field Team)

### Inner Setting

At the inner setting, we identified several challenges, including fragmented communication between implementers and authorities. Conflicting leadership roles led to mixed messages during the event, exacerbated by a lack of clear (and overall) leadership. Additionally, implementers and authorities did not conduct post-event evaluations, hindering a thorough assessment of the campaign’s effectiveness or opportunities for making improvements for future campaigns. The event also faced shortages of supplies and equipment, impeding the smooth execution of vaccination efforts.

Regarding the work infrastructure, implementers and authorities mentioned the need for more clarity regarding the vaccination strategy and more coordination at the vaccination point. This was confirmed in the field notes:

> “The initial order was that they had to cover the entire selected area and, in the end, they were left at a fixed point (which was later changed back to mobile). Another case was with a collaborator – the initial directive was house to house and then [it was] changed to fixed, but without communicating [to the population] with the megaphone” (MDVC Implementer)

> “…there was a lack of coordination from the beginning. There were slips of paper with the distribution of the places. They mentioned that they would deliver maps of the vaccination sites, and in the end, they did not deliver anything” (Research field Team)

Regarding materials and equipment, the three study groups—community, MDVC implementers, and research field team—reported a shortage of supplies:

> “Even though it was 11 a.m. there were no more vaccines. They had been there since 9 a.m., that is to say, just 2 hours…” (Urban Community Member)

> “Those in charge of promotion were very upset because last year [the Ministry of Health] offered various materials such as flyers, radio spots, brochures, and to date, they have not been fulfilled” (Research field team)

> “… [There was an] inadequate supply of PPE and there were missing collars to identify the vaccinated dogs” (MDVC Implementer)

Moreover, MDVC implementers, authorities, and the research field team noted the need for better training among vaccination teams; not only in vaccinating, but also in dealing with people:

> “During the vaccination, there were 3 brigades that crossed each other in the same block in different areas due to a misinterpretation of the maps from some brigades and supervisors” (MDVC Implementer)

> “At the time of vaccination, the thermos lid was left open” (Research field Team)

> “Many of the vaccinators from the army did not know how to vaccinate or how to fill out the card or records; many had been replaced at the last minute by untrained people” (Urban Community Member)

> “It would be great if it were a veterinarian, wouldn’t it? It would give me more confidence (…). [If there were a veterinarian] people would say to each other ‘It was a veterinarian who is vaccinating’. Then, everyone would spread the word around” (Urban Community Member)

> “At the beginning, they do not tell you what they will give the dog. They do not tell you: ‘good afternoon/good morning, we are going to give him this medicine, it is called. I don’t know - Barrovirus. It is to prevent this disease…’ So sometimes you are afraid to bring your dogs to them” (Urban Community Member)

### Individuals

At the individual level, beliefs about rabies vaccination can pose significant barriers. For instance, periurban participants mentioned that some neighbors think that *‘this vaccine should last for more years, so they will not need to be vaccinated again*’. They also mentioned that some think vaccinating older dogs is unnecessary or risky, fearing it might make the dog sick. Also, urban participants mentioned that there is a general lack of knowledge about rabies and that everybody, including veterinarians, *“always give priority to distemper, to parasites, and do not talk about rabies. [The vet] does not tell you that you have to vaccinate your dog [against rabies]*”

Implementers, authorities, and community members also reported that while companion dogs (pets) are usually considered part of the family and are vaccinated, guard dogs are not and are thus less likely to be vaccinated. They also perceive that most vaccinated dogs are purebred, and that these dogs are at lower risk than most dogs because they are kept indoors.

> “Dogs that are vaccinated against rabies are also considered pets. But dogs that are not, are dogs that guard the house, the stable dog, the watchman’s dog. They are considered as working dogs, community dogs, and not as pets” (MDVC Authority)

Implementers and authorities also emphasized the need for community agents to help raise public awareness at both community and individual levels.

Another important point was the perception of economic access to the campaign, as the community needed to find out if the MDVC was free or not. Some people mentioned that “*it was a relief that the campaign was free last year” (Periurban Community member)*.

The COVID-19 pandemic impacted MDVC participation at the individual and outer setting domains. Within the individual domain, the pandemic has affected households and the community in many ways. Participants, particularly from peri-urban areas, described being financially affected by the pandemic and the need to work on weekends—the usual time for MDVCs—to compensate for lost income. This represents a significant lifestyle change from pre-pandemic times. Initially, strict military-enforced lockdowns even prohibited families from walking their dogs outside, confining dogs indoors with their families. Consequently, many participants reported some dog ownership behavior changes post-pandemic, specifically, walking their dogs less frequently than before the pandemic:

> Woman, urban area: “In my house, since I have my children, what I don’t do during the week, I have to do on the weekend, go to the market. Not only that, but I mean, yes, my life has changed after Covid” Facilitator: “Tell me more”

> Woman, urban area: “Because… I have become very, like… I don’t want anyone to come into my house with dirty shoes. [With] my dog, it is the same, I don’t take her out much, because of COVID, I take her every month to where they bathe her, to the vet […] From the house she walks with her little legs and with the leash. But then [after the bath], completely clean from the vet […], we carry her and we leave her in the house. […] I am, a little bit like that, nervous for me, for my little [son]”

Participants also mentioned spending more time indoors post-pandemic, resulting in being less informed about community events through informal conversations amongst neighbors and perceiving a lower risk of their dogs contracting rabies than before.

### Outer Setting

For this domain, we considered governmental and academic strategic partnerships affiliated with the MDVC intervention: 1) the Ministry of Health, 2) the Universidad Peruana Cayetano Heredia (UPCH), 3) others (municipalities, brigade chief, volunteers, etc.), and 4) private entities (e.g. veterinary clinics, political groups).

Authorities and implementers noted problems with competing events, such as that of a local political campaign offering free deworming on the same day as the MDVC at the stadium: *“Dogs were being dewormed at the stadium by the FUERZA AREQUIPA campaign” (MDVC Implementer)*. They reported that overlapping health campaigns create confusion, divide public attention, and reduce the impact of the MDVC by diverting resources and participants’ focus, ultimately undermining vaccination efforts. Competing events not only create confusion for participants, but also impact the coordination of planned activities. For instance, the implementers reported that municipalities provide support to the MDVC by sending vans and pick-up trucks, typically used for public security, for transporting vaccination teams. However, “*this is often impossible because, on the day of MDVC, they have to support other events– celebrations, fairs, etc.*” *(MDVC Implementer)*.

Community participants also reported being confused by private veterinarians who show up at the communities a day or two before the MDVC to vaccinate, thinking the private veterinarians are part of the MDVC, or being confused by their claims that government-subsidized vaccines were of poor quality. This was confirmed by authorities and implementers who reported that these private veterinarians would arrive in the district before the MDVC and charge individuals for the vaccines or provide the rabies vaccine for free but charge for a related service such as deworming.

> “There is a man who comes and charges you. Sometimes you get the rabies vaccine for free, but sometimes you have to pay” (Periurban Community Member)

> “…a [private] veterinarian one day told me not to have my dog vaccinated at the MDVC because as it [vaccine] comes from the state, there is no guarantee [about its quality]. The veterinarian was giving the vaccine from a certified company and that’s why it’s not free” (Urban Community Member)”

> “A person told us that most of the neighbors were in a meeting. She remarked that it would have been helpful to receive advance notice so that no other events would be scheduled for that day” (Research field team)

MDVC implementers and authorities commented that advertisements through megaphones should be mobile and not limited to just the day of the MDVC. They also suggested holding a press conference to inform the community about the MDVC sites ahead of time.

Furthermore, community, implementers and authorities agreed that the people would rather go to the COVID-19 vaccination campaigns than the rabies campaign, because COVID-19 forced them to take care of their family first, and put their dogs’ health on the last place.

> “There are some people who, …that think that way, prioritize only their children, and then the animals, the animals are left on the streets” (Periurban Community Member)

Periurban and urban participants asked for better coordination between municipalities or authorities so that there is no crossover between rabies vaccination and COVID-19 vaccination.

Several community members mentioned that municipal health representatives should conduct regular health education about dog ownership and the importance of pet vaccination against rabies, since the media recently has mostly been focused on COVID-19, with no information about rabies:

> “I also think that today that it is almost three years since we are with the pandemic that more interest has been given to covid and that rabies has been neglected” (Urban Community Member)

### Innovation: MDVC

The implementers and authorities held several meetings to design the MDVC, specifically setting up vaccination points and methods, establishing purchase and operating costs, preparing equipment, and recruiting vaccinators and field teams. The research field team registered the coordination details:

> “Currently, all the microreds [health networks] are in the preparatory phase of their action plans for their ‘VANCAN’ [local name of MDVC]; the dates will vary according to the availability of vaccines” (MDVC Authority)

Some implementers showed interest in improving their planning strategies. Initially, they used paper maps to select vaccination points without considering distance, often choosing inconvenient locations. To improve, some started using optimization points^11^ provided by the UPCH team, while others stuck to the traditional method of revisiting previous vaccination sites.

Implementers identified several barriers to achieving vaccination coverage, with the most significant being a limited budget shared with other vector-borne and zoonotic disease programs. They also reported being stretched thin across multiple activities and struggled to manage their responsibilities effectively.

> “One of the problems is that at the regional level, the issue of rabies is not prioritized, right? It is not a priority for the regional government, but if the regional government would take interest in the problem, it would be reflected in the actions that they take normatively and also budgetarily. As they say, ‘unfortunately there are not enough resources to be able to carry out actions to control rabies in Arequipa, which is not a priority’” (MDVC Authorities)

> “They said that they had not had support from the local government and, unlike other years, they were just finishing the management of the previous mayors, there was very little budget” (Research field Team)

## Discussion

In the midst of a dog rabies epidemic, proper implementation of any strategy is urgent for multiple reasons. First, the risk of human rabies cases is imminent, highlighted by an already reported human death from rabies^32^, the high rate of dog bites, and the low proportion of medically attended bites^21^. Second, authorities might replace an inadequately executed strategy with another that, if also poorly implemented, could worsen the epidemic. Third, poor execution may foster distrust towards health authorities, potentially decreasing public participation in future initiatives.

We examined the decrease in MDVC coverage during a canine rabies epidemic in Arequipa, using the CFIR to ensure a comprehensive lens to analyze system challenges (and the nuanced interplay between these) and to structure the findings. Despite scientific evidence supporting the use of fixed points as the main strategy for MDVCs, this study found significant challenges in all domains resulting in an inadequate implementation of the campaigns, limiting its coverage and overall effectiveness. Key issues included frequent and serious deviations from pre-established MDVC plans, poor communication about vaccination schedules and locations, inadequate training for vaccination teams, and logistical problems like insufficient supplies and transportation. A critical problem that is both the result and root of the previous issues is the inconsistent location of the fixed vaccination points. Moreover, competing health and municipal events, misinformation or competition from private veterinarians, and lasting effects from the COVID-19 pandemic have all also affected the MDVC coverage. Community members also reported barriers that could be construed as vaccine hesitancy.

The literature on vaccine hesitancy emphasizes the “3 Cs” model^33^, confidence (trust in vaccine efficacy and safety), complacency (low perception of risk), and communication (information on vaccines and the campaigns). We found that confidence was often undermined by misinformation or conflicting messages from private veterinarians. Complacency appeared in the form of underestimating rabies risk, especially regarding which dogs to vaccinate (pedigree vs. guard dogs) or how often vaccinations were truly needed, leading some to skip current campaigns. Lastly, communication was a pivotal factor in Arequipa; inadequate dissemination of information regarding vaccination schedules and locations significantly hampered community participation. Addressing these three elements is crucial for improving vaccine uptake and the overall success of future campaigns.

Currently, there is a shortage of personnel for the MDVC in Arequipa. The involvement of ‘lay dog vaccinators’ (individuals trained as vaccinators but not veterinarians or health professionals) can mitigate the effect of this shortage in resource-limited settings during a mass campaign^34^. However, concerns about their training can deter community participation, as reported above. To build public confidence and ensure an adequate vaccination process, clear policy guidelines are needed regarding selecting, training, certifying, deploying, and monitoring these vaccinators^34^. Importantly, incorporating private veterinarians into the vaccination strategy could both, turn them into valuable allies, and instill trust among dog owners^8,34^. Participation of private veterinarians could involve ‘lay dog vaccinators’ training and monitoring, using their local connections and position as health leaders to promote the MDVC, and providing veterinary services alongside public campaigns.

Reducing complacency about rabies risk is crucial for improving vaccination coverage. Our findings align with studies showing that communities in endemic areas often underestimate the true risk of rabies transmission to humans, despite its high case fatality rate^35–37^. This is further exacerbated by the epidemiological silence surrounding human rabies cases, as the absence of recent cases may contribute to a false sense of security which can delay timely response and vaccination efforts, increasing the risk of outbreaks. Research has demonstrated that even brief lapses in mass dog vaccination can lead to outbreaks, as seen in rabies-endemic regions of Latin America and Asia^3,12,14,38,39^. Furthermore, there is also a common misconception that rabies is a disease that only affects dogs, overlooking the potential impact on human health. This problem can be reduced by raising awareness about the true risks of rabies to both humans and animals, emphasizing that all dogs, regardless of breed, purpose or history of vaccination need to be vaccinated regularly^40–43^.

Effective communication and health promotion are crucial for ensuring communities are well-informed about vaccination campaigns, allowing time for preparation and increasing participation^44,45^. Clear leadership and coordination within the implementation team are equally important to ensure schedules are followed. Addressing logistical challenges, such as adjusting vaccination times to fit community needs^46,47^ and optimizing the location of vaccination sites^10,11^, can further boost coverage. Campaign visibility could also be enhanced, especially in remote areas, through strategies like loudspeakers on garbage trucks to reach difficult-to-access locations^48^ and promotional materials being prominently displayed in public spaces like markets, schools, bus stops, parks, and churches. Involving the community in planning and decision-making, along with using culturally relevant communication strategies, can significantly boost participation and trust^44,49^. Engaging local leaders, volunteers, and youth will be key to fostering community ownership for vaccination campaigns, a strategy that has worked well for other rabies and other immunization programs^49–52^. Addressing misconceptions about rabies and the effectiveness of vaccines through targeted education campaigns is critical^53^.

We emphasize the critical need to analyze the factors that influence the planning, implementation, and effectiveness of mass vaccination campaigns. To ensure success in dog rabies elimination, future research must prioritize the evaluation of innovative approaches and alternative delivery methods that effectively reach underserved populations^54^ and motivate dog owners to commit to annual vaccination of all of their dogs. The impact of integrating additional services to MDVCs, such as deworming or sterilization should also be examined^55–57^. It is important to investigate whether year-round vaccination in high-risk areas^58,59^, for example, communities with high dog population turnover, could ensure sustained coverage. Expanding the scope of research to include the efficacy and effectiveness of various One Health strategies—addressing dog rabies vaccination, responsible dog ownership, and accurate dissemination of MDVC information and increasing community engagement—will be pivotal in refining public health interventions and achieving the global goal of eliminating human rabies deaths.

Our study has some limitations; we left room for future exploration by not delving into participants’ perceptions of COVID-19 vaccination, thus leaving the correlation between attitudes towards COVID-19 vaccination and participation in MDVCs open for further research. Additionally, we did not involve authorities from the central offices of the National Institute of Health and the Ministry of Health in Lima, where official guidelines and procedures are established. Moreover, the retrospective nature of the qualitative data calls for cautious interpretation and consideration of the temporal aspects when drawing conclusions. However, our study’s strengths include a diverse and representative sample, engaging community members from both urban and periurban areas, detailed field notes, and a collaborative workshop with MDVC implementers and authorities. Utilizing the CFIR framework in our analysis facilitated the co-creation of actionable strategies. This comprehensive approach provides a deep understanding of post-COVID-19 MDVC participation decline and yields practical community-based recommendations.

## Conclusions

Our study offers valuable insights into the decline in MDVC vaccine coverage, a problem that already existed and was further exacerbated by the COVID-19 pandemic. By examining perspectives from diverse stakeholders, and collaborating closely with MDVC implementers and authorities, we identified key factors influencing participation rates and impeding reaching herd immunity. The application of the CFIR framework enabled a thorough analysis of system components and challenges, leading to actionable recommendations that consider the unique contextual challenges posed by the pandemic and varying socio-economic conditions. We uncover that implementing evidence-based, data-driven solutions for MDVCs remains a novel approach, where resistance or skepticism toward new strategies is still prevalent. Our findings also underscore the importance of addressing community-specific barriers and enhancing communication and confidence between health authorities and the public. The study highlights the need for tailored interventions that are responsive to the evolving landscape of public health needs and the socio-economic realities within communities. Continuous monitoring and evaluation, incorporating both qualitative and quantitative methods, are crucial for the ongoing improvement of MDVC. While our study includes a group model building workshop, it is essential to maintain a reflective and skill-building process to sustain progress. Future One Health research and policy efforts could build on these findings to enhance the effectiveness and sustainability of MDVCs in diverse settings.

## Other Information

### Ethics approval and consent to participate

Institutional Review Board approval was obtained for the focus groups and the workshop from Universidad Peruana Cayetano Heredia (approval identification numbers: 65369 and 207735), Tulane University (approval identification numbers: 14–606720 and 00000339), and the University of Pennsylvania (approval identification numbers: 823736 and 850695). All participants gave written consent to participate and to be audiotaped.

### Availability of data and materials

The datasets generated during and/or analyzed during the current study are available from the corresponding author upon reasonable request.

### Authors’ contributions

RCN: Conceptualization (lead); supervision (supporting); writing – original draft (lead); writing – review and editing (equal); methodology (supporting); formal analysis (supporting); funding acquisition (lead).

LOC: Writing – original draft (supporting); writing – review and editing (equal)

JC: Investigation (supporting); data curation (lead).

EWD: Investigation (supporting)

LDT: Investigation (supporting); writing – review and editing (equal)

GP: Investigation (supporting)

SER: Investigation (supporting); writing – review and editing (equal).

VPS: Conceptualization (supporting); supervision (lead); writing – original draft (supporting); writing – review and editing (equal); methodology (lead); formal analysis (lead).

## Data Availability

All data have been anonymized and uploaded to the submission system solely for the purposes of editorial review. Data will be made available upon reasonable request to Ricardo Castillo-Neyra, the corresponding author. Please note that participants signed a consent form stating that their data would be kept confidential. Therefore, we are cautious about making the data fully publicly available.

## Acknowledgments

We gratefully acknowledge the Gerencia Regional de Salud de Arequipa and Red de Salud Arequipa Caylloma for their work to eliminate dog-mediated human rabies from Arequipa. We thank Antuanette Vela and Cinthia Mamani for organizing the workshop, John Quispe for organizing the focus groups, Liz Frisancho for conducting the transcripts, and Marianne Luyo and Joanna Brown for developing the codebook.

